# Virus neutralizing antibody responses after two doses of BBIBP-CorV (Sinopharm, Beijing CNBG) vaccine

**DOI:** 10.1101/2021.07.15.21260362

**Authors:** Tamás Ferenci, Balázs Sarkadi

**Author notes:** Name and address of the corresponding author: Balázs Sarkadi, Magyar Tudósok krt. 2, 1117 Budapest, Hungary.

## Abstract

**Background:** Limited information is available on the effectiveness of the BBIBP-CorV (Sinopharm, Beijing CNBG) vaccine, especially in the elderly, despite the fact that it is approved in more than 50 countries.

**Methods:** Virus neutralizing antibody titres, as a rapidly available but highly predictive surrogate marker, were measured after two doses of the BBIBP-CorV vaccine in 450 subjects. Results were analyzed in a multivariable model accounting for age, sex and time since the administration of the second dose of the vaccine.

**Findings:** Sex and time since the second dose had little association with the antibody titres. Age, however, was highly relevant: measurable antibody levels were present in about 90% of individuals below the age of 50, but antibody production after BBIBP-CorV vaccination was strongly reduced with increasing age. A large number of elderly subjects, reaching 25% at 60 years, and up to 50% at ages over 80, were found not to produce any protective antibody.

**Interpretation:** Neutralizing antibody titre, as a correlate of protection for COVID-19 disease susceptibility, should help to evaluate the effectiveness of the BBIBP-CorV vaccine. Results suggest that proper measures should be undertaken to prevent a potential outbreak of COVID-19 in BBIBP-CorV vaccinated but eventually unprotected elderly individuals.

**Funding:** No specific funding was used to carry out the study.

**Research in context:** *Evidence before this study:* The BBIBP-CorV (Sinopharm, Beijing CNBG) vaccine was found to be effective, but the Phase 3 trial included few women, almost no elderly subject and no subject with preexisting medical condition. It is therefore especially important to monitor the real-world effectiveness of this vaccine, however when searching Pubmed for (Sinopharm OR BBIBP-CorV) AND vaccine AND (effectiveness OR efficacy OR antibody OR titre) in text words with no language or date restriction and with translation of the terms enabled, we found no such published study. The most rapid way to generate such evidence is the assessment of antibody response which was found to be highly predictive for disease susceptibility. This is of crucial importance as the vaccine is approved in more than 50 countries, and hundreds of millions have already been vaccinated with many planned to be vaccinated in the near future with this vaccine.

*Added value of this study:* Virus neutralizing antibody titres were measured after two doses of the BBIBP-CorV vaccine in 450 subjects. Results were analyzed in a multivariable model accounting for age, sex and time since the administration of the second dose of the vaccine. We found that sex and time since the second dose had little association with the antibody titres. Age, however, was highly relevant: while the vast majority of those below 50 years had measurable antibody levels, this rate quickly dropped with increasing age. In subjects aged 60 years, quarter, in those above 80 almost half had no detectable neutralizing antibody.

*Implications of all the available evidence:* The real-world effectiveness of the BBIBP-CorV vaccine should be carefully monitored. Elderly subjects are especially prone to produce no protective antibody after vaccination. This should be monitored, and proper measures should be undertaken to prevent a potential outbreak of COVID-19 in BBIBP-CorV vaccinated but eventually unprotected elderly individuals.

## Introduction

The use of the BBIBP-CorV (Sinopharm, Beijing CNBG) vaccine in Hungary was authorized by the Hungarian national drug and food evaluation authority, based on a governmental decree in January^1^. At the time of this authorization there were no publicly available Phase 3 evaluation data, and unpublished data partially released by the Hungarian authorities suggested that limited information was available about the use of the Sinopharm vaccine, especially in elderly people: these interim data reported that 99.3% of the subjects in Phase 3 was below 60^2^. As of 27 June 2021, 1.05 million people received the first dose with this vaccine in Hungary, and 1 million received the second dose too^3^. Of these, more than half (54.1% from those receiving the first dose) were above the age of 60^3^. Since no other EU countries authorized the use of Sinopharm vaccine, it is especially important to follow the potential protective effect of this vaccination in an EU member country, focusing on elderly people.

Correlates of protection^4^ for SARS-CoV-2 are not yet firmly established^5^, but a growing body of evidence suggests that neutralizing antibody levels are highly predictive for protection against symptomatic disease^6–13^. While very limited empirical evidence exists specifically about the Sinopharm vaccine in this respect, traditional immunology knowledge and experience with prior vaccines suggest that the technology of the Sinopharm vaccine (whole inactivated virion with alum adjuvant) results in a Th2-skewed immune response^14,15^. This also suggests that in case of the Sinopharm vaccine, antibody response may be a good correlate, which is further reinforced by the fact that in case of a technologically very similar SARS-CoV-2 vaccine, CoronaVac, the Phase 2 study failed to find clear evidence of cellular response using ELISpot^16^. (Such study was unfortunately not carried out during the Phase 2 of the Sinopharm vaccine^17^.)

On May 24, 2021, a report on the Phase 3 trial of the Sinopharm vaccine, showing positive efficacy and safety data, was finally published^18^. However, the paper revealed that 84.4% of the trial participants were male, 98.4% were aged less than 60 years and 100% were healthy (as it was an inclusion criteria, with “healthy” being investigator-judged with no precise definition). This is in stark contrast with the trials of vaccines that are authorized in other EU countries^19^, for instance, in the Phase 3 trial of the BNT162b2 vaccine (Comirnaty, Pfizer/BioNTech) 42.2% of the subjects was above 55 years of age^20^ and 20.3% had one or more underlying disease^21^. As a result, there is very little direct evidence available about the efficacy and safety of Sinopharm vaccination among women, in elderly, and those with underlying diseases, thus a study in these regards is highly important, especially as these are also largely the risk groups for SARS-CoV-2 infection.

In addition, news reports from the United Arab Emirates, Serbia and other countries using mostly the Sinopharm vaccine indicated limited protection against SARS-CoV-2 virus infection in elderly people after vaccination with Sinopharm. Anecdotal cases from Hungary also pointed to possible inefficient protection by even two doses of Sinopharm vaccine: several hundreds of such test results were informally reported in the press and social media.

Therefore, a proper analysis of virus neutralizing antibody measurements should have a major importance in evaluating the potential efficacy of Sinopharm in elderly, thus helping to prevent a potential outbreak of COVID-19 in vaccinated but eventually unprotected individuals. This question is crucial as more than 50 countries approved the Sinopharm vaccine as of late June, 2021^22^ and it is on the World Health Organization’s Emergency Use Listing^23^.

## Methods

Antibody measurements (initiated by the vaccinated individuals) after at least 14 days of obtaining the second vaccine dose of the BBIBP-CorV (Sinopharm, Beijing CNBG) vaccine were collected from >18 year old subjects. Known prior SARS-CoV-2 or current infection were exclusion criteria, but subjects were otherwise unselected. Information on the age and sex of the subjects and the time past after the second dose were also collected. All subjects signed a full written informed consent allowing the later publishing and anonymous statistical analysis of the collected data. De-identified data published based on this consent were openly available to the public before the initiation of the study, and can be found – along with the full analysis script – at https://github.com/tamas-ferenci/SinopharmAntibodyResponse.

After obtaining venous blood samples from the individuals, the virus neutralizing antibody measurements were performed in an accredited laboratory using an FDA emergency use approved and internationally accredited method (ADVIA Centaur SARS-CoV-2 IgG, COV2G), shown to correlate with direct virus neutralizing tests^24^. For a comparison, we have also obtained anonymous neutralizing antibody laboratory data for 45 individuals, vaccinated by two doses of the Pfizer/BioNTech RNA-based vaccine. Neutralizing antibody titres provided by the applied antibody assay are measured in the “Centaur index” (Ci) arbitrary unit and are categorized according to the manufacturer as follows:

less than 1 Ci: no protective antibody present,

1-10 Ci: moderate virus neutralizing antibody titre,

10-30 Ci: strong virus neutralizing antibody titre,

30-150 Ci: outstandingly strong virus neutralizing antibody titre.

To investigate the effects of age, sex and time after the second dose, a multivariable model was used in which these variables were entered as covariates, with the response being the titre. Age and time after the second dose were spline-expanded with thin plate regression splines to allow for a potentially non-linear effect^25^. Given the high number of zero titre measurements and the highly skewed nature of the non-zero measurements, a hurdle-lognormal model was used, with the same formula for the hurdle. As an alternative analysis, standard logistic regression was applied with the titre dichotomized at 1 Ci cutoff (no protective antibody), with the same covariates and similar spline expansion. In case of the Pfizer/BioNTech vaccine, antibody titres were measured exactly 28 days after the second dose in all subjects, so time since the second dose was not included in these investigations, but the models were otherwise the same as for the Sinopharm data.

Statistical analysis was carried out under the R statistical environment version 4.1.0^26^ using package brms version 2.15.0^27^. Models were estimated in a Bayesian approach with default weakly informative priors of brms applying the Markov Chain Monte Carlo (MCMC) method using 4 chains and 2000 iterations with 1000 warmup^27^. Convergence was assessed with the potential scale reduction factor (Gelman-Rubin statistic) Rhat.

Results are presented as predicted probabilities that the titre is below a certain threshold, which was calculated by obtaining 4000 posterior samples of the linear predictor and then the probability was manually calculated taking the hurdle into account. The sample size for the Sinopharm vaccinated subjects was large enough so that the estimates are sufficiently precise, as evidenced by the width of the credible interval.

## Results

Within the period of April 1 – June 21, 2021, 497 measurements were performed in Sinopharm-vaccinated subjects. Of these, 6 had no information on the date of the second dose and 1 had missing data on age; of the remaining 490 measurements, 40 was carried out within 14 days of the second dose. Excluding these resulted in a final dataset with n=450 subjects that was analyzed.

Figure 1 shows the age distribution of the individuals, both male and female, participating in the antibody measurements. Since these were measurements initiated by private persons, mostly elderly individuals requested the antibody measurements, and the peaks of the curves are about the ages 65-75 for both sexes. Still, data from numerous younger and older individuals are also included in the present analysis. Of the participants, 208 (46·2%) were male.

**Figure 1.**
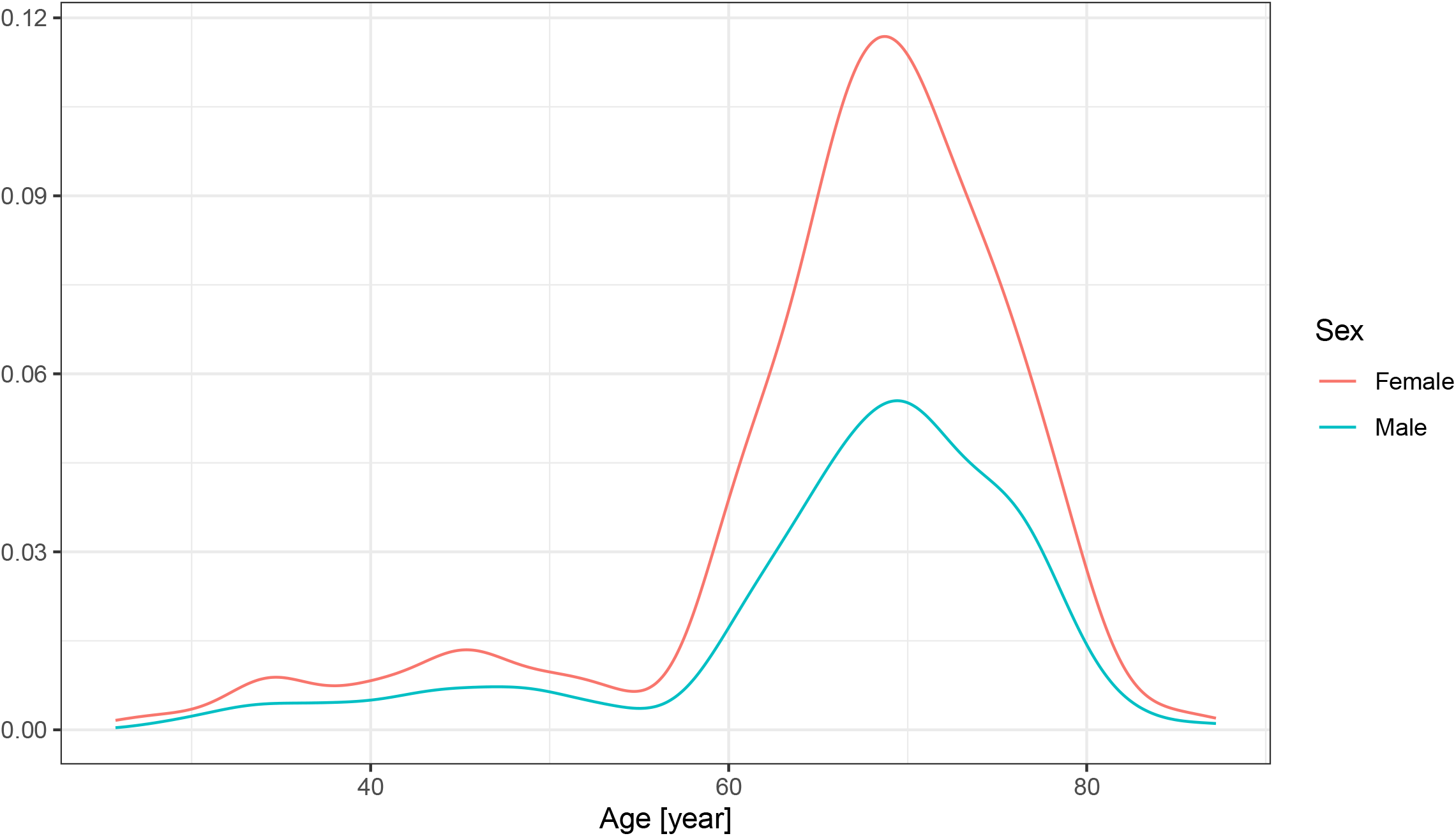
Age distribution of individuals for virus-neutralizing antibody measurements after two doses of Sinopharm vaccine.

The median time after the second dose was 23 days (lower quartile: 18 days, upper quartile 30 days).

As shown in Figure 2, the overall antibody titre showed a decreasing tendency by the increasing age, moreover, a large number of subjects with no antibody (0 titre) was present in the elderly population.

**Figure 2.**
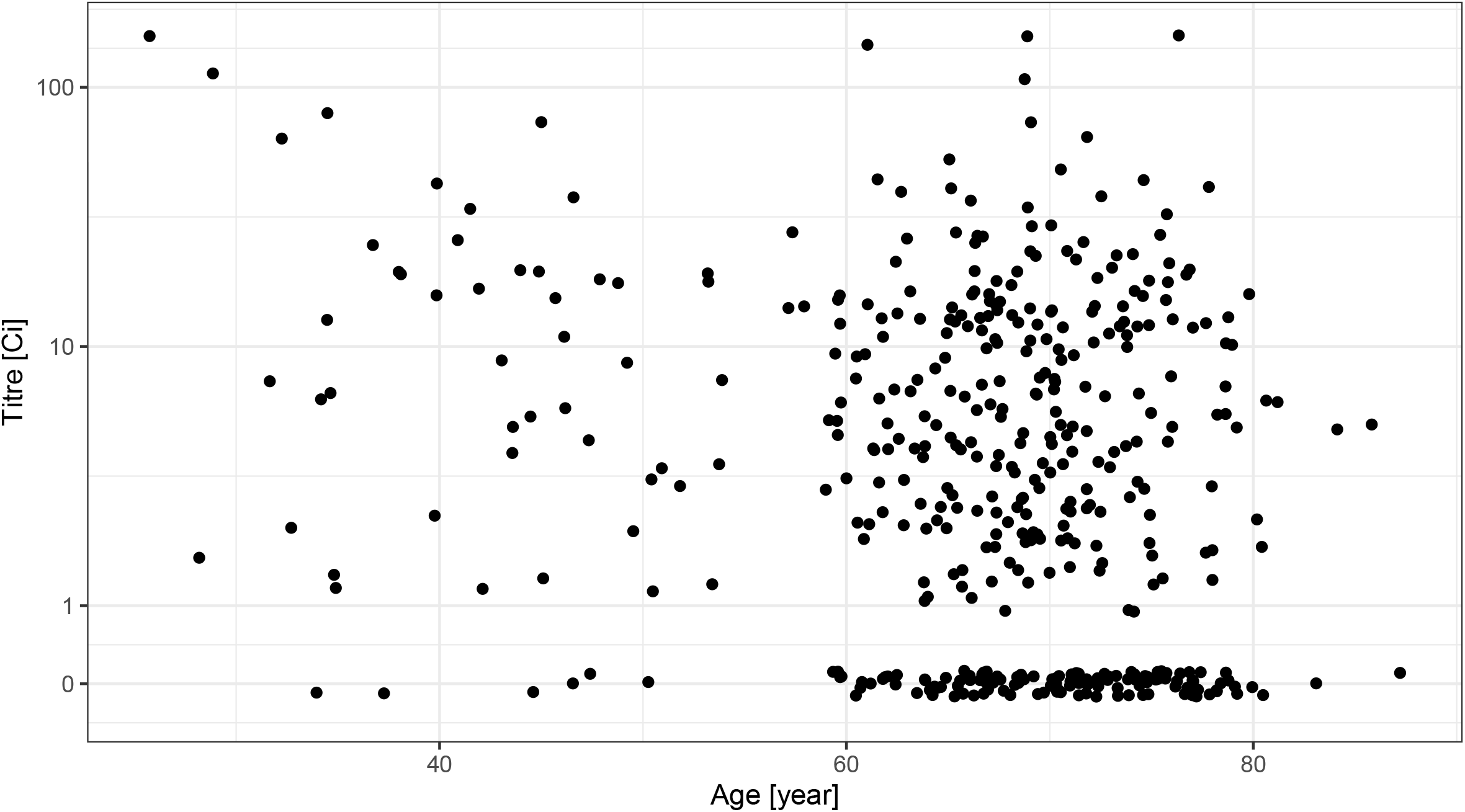
Distribution of the virus neutralizing antibody titre according to the age of the subjects after two doses of the Sinopharm vaccine shown as jittered scatterplot. (Data are shown in pseudo-log scale.)

The distribution of the measured antibody titre showed no apparent univariate association either with the number of days since the second dose (Figure 3), or with the sex of the subjects (Figure 4).

**Figure 3.**
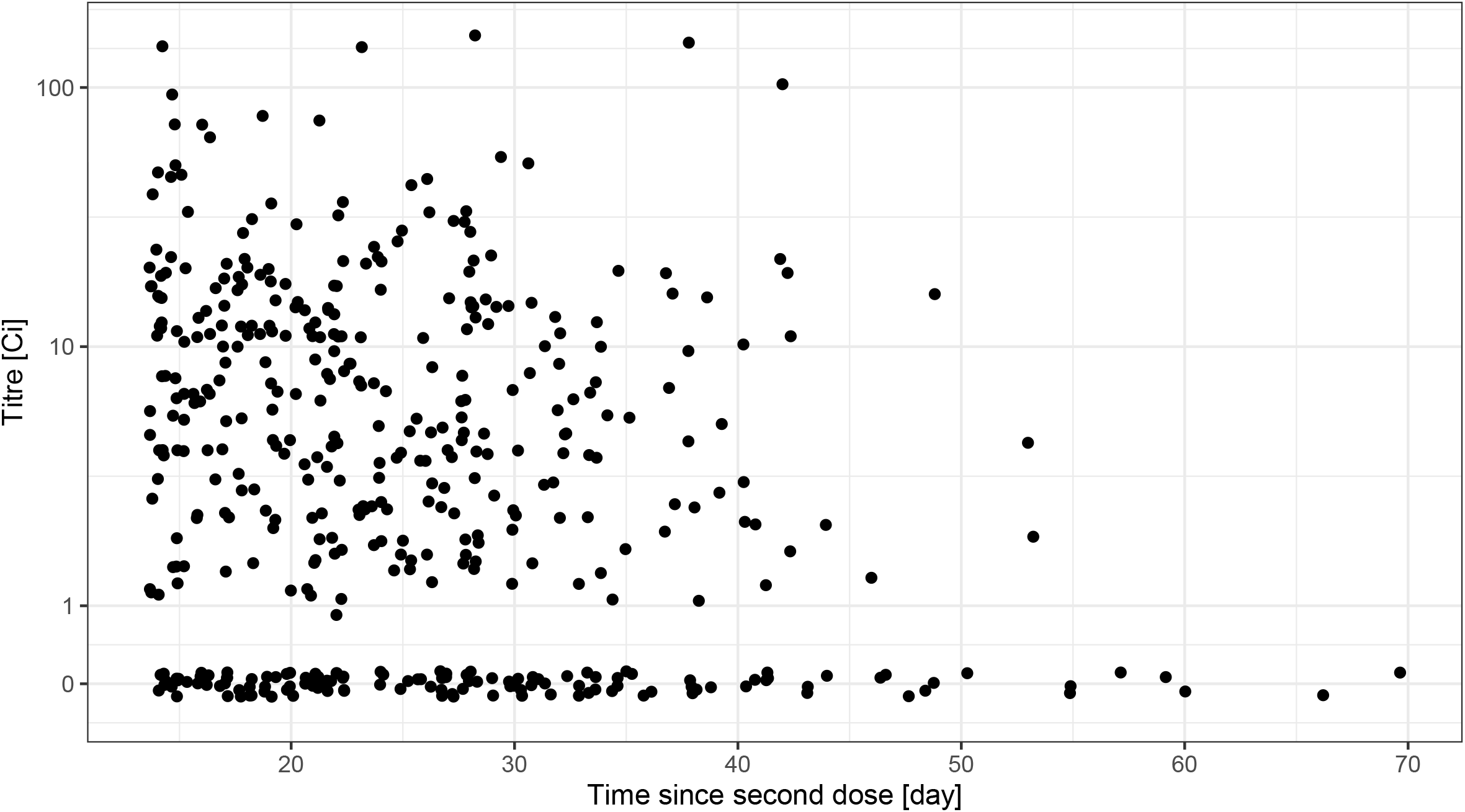
Distribution of the virus neutralizing antibody titre according to the time-period of the antibody measurement following the vaccination by two doses of the Sinopharm vaccine shown as jittered scatterplot. (Data are shown in pseudo-log scale.)

**Figure 4.**
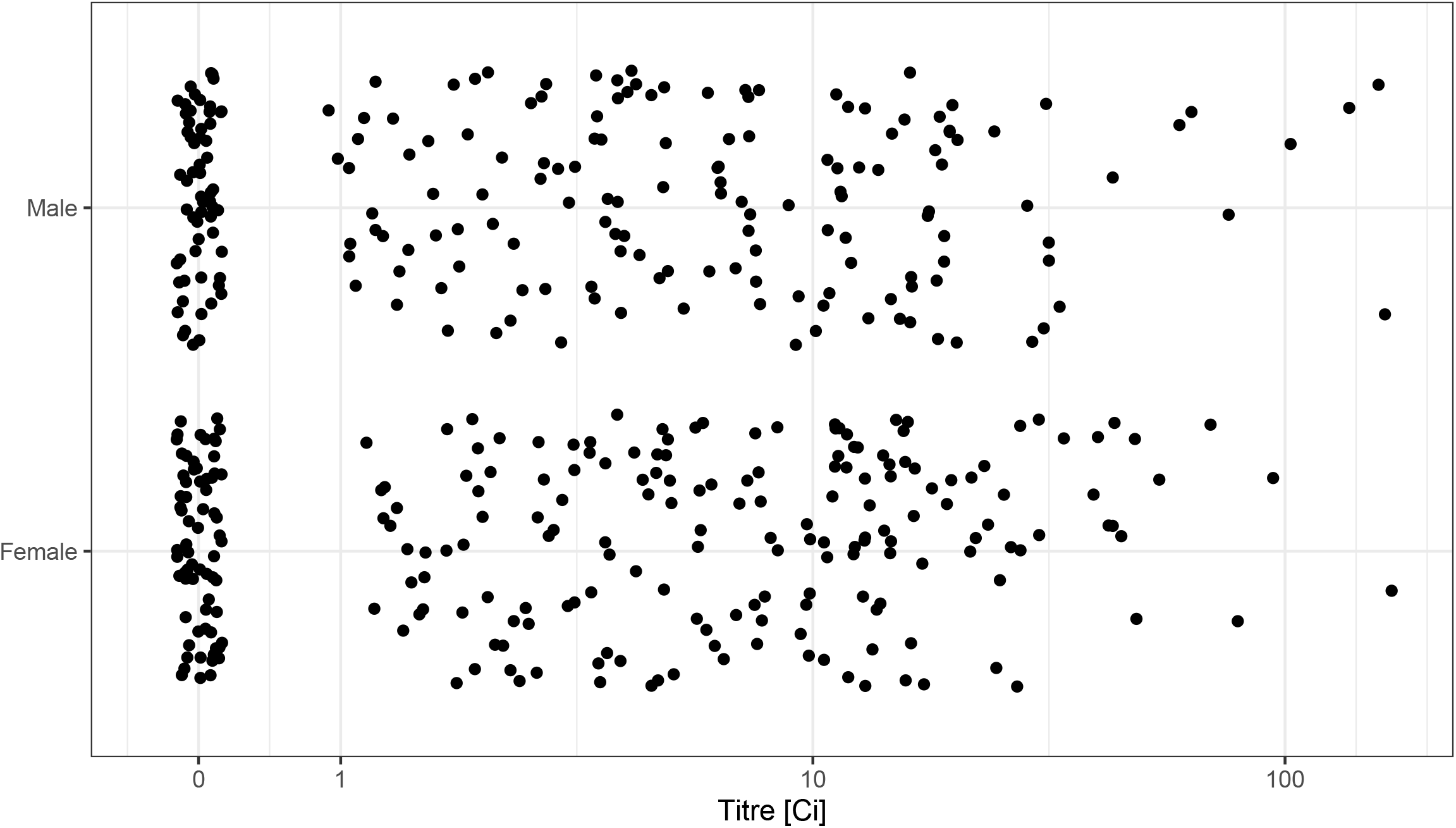
Distribution of the virus neutralizing antibody titre according to the sex of the subjects after two doses of the Sinopharm vaccine. (Data are shown in pseudo-log scale.)

In the following, we performed a multivariable modelling to estimate the probability of having low neutralizing antibody titre after obtaining two doses of the Sinopharm vaccine. Rhat was 1.00 for all parameters, indicating convergence.

As shown in Figure 5, the increasing age of the subjects shows a significant correlation with the lack of antibody production (titre below 1, no response), or low antibody titre (titre below 10, weak response). At about the age of 60 years, the estimated probability of the lack of antibody response was about 25%, and this value increased up to about 50% at the age of 80 years. Regarding the expected low titre response, at the age of 60 years this reached 60-75%, further increasing with increasing age.

**Figure 5.**
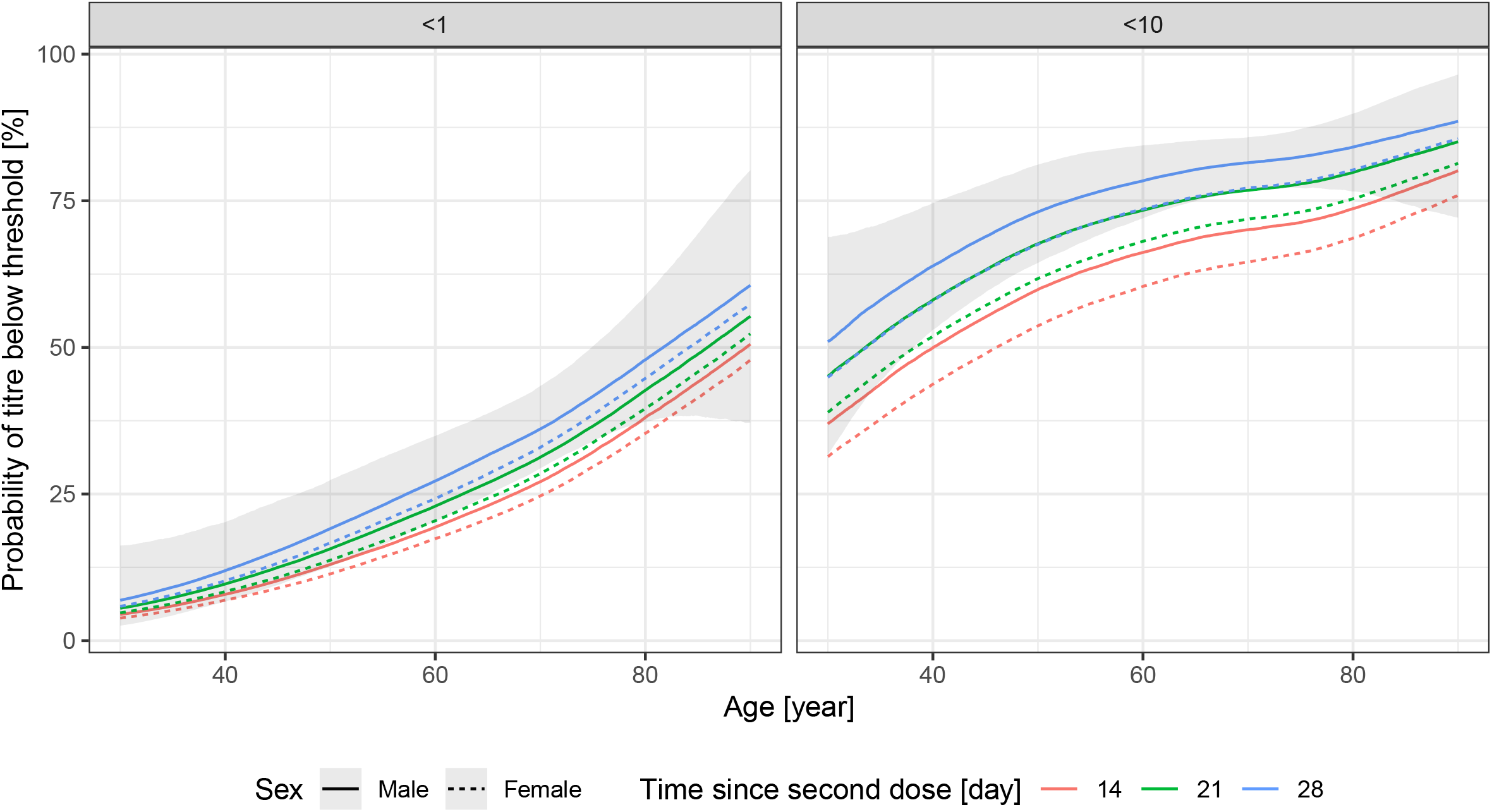
Effects of the age and the sex of the subject, and the time-period between the vaccination and the measurement on the probability of the lack of antibody production (titre below 1, left panel) and of low antibody titre (titre below 10, right panel) after two doses of the Sinopharm vaccine. 90% credible interval is shown for males, 28 days post second dose.

The number of days between the vaccination and the antibody measurement had little negative effect, while in the case of female subjects the estimated probability for the lack of antibody production or lower antibody titre was somewhat smaller.

Logistic regression analysis confirmed these findings (data not shown here).

Although we have data only for a relatively small number (45) of subjects vaccinated by the Pfizer/BioNTech vaccine, these (Figure 6) indicate a drastically lower probability of having low, and especially zero antibody titre in the vaccinated individuals. The increasing age also had a negative effect on the antibody titre in the case of the Pfizer/BioNTech vaccine, but no subject was found in this group producing no protective antibody (that is, a titre below 1), the probability of which was estimated to be less than 10% even in the oldest age group.

**Figure 6.**
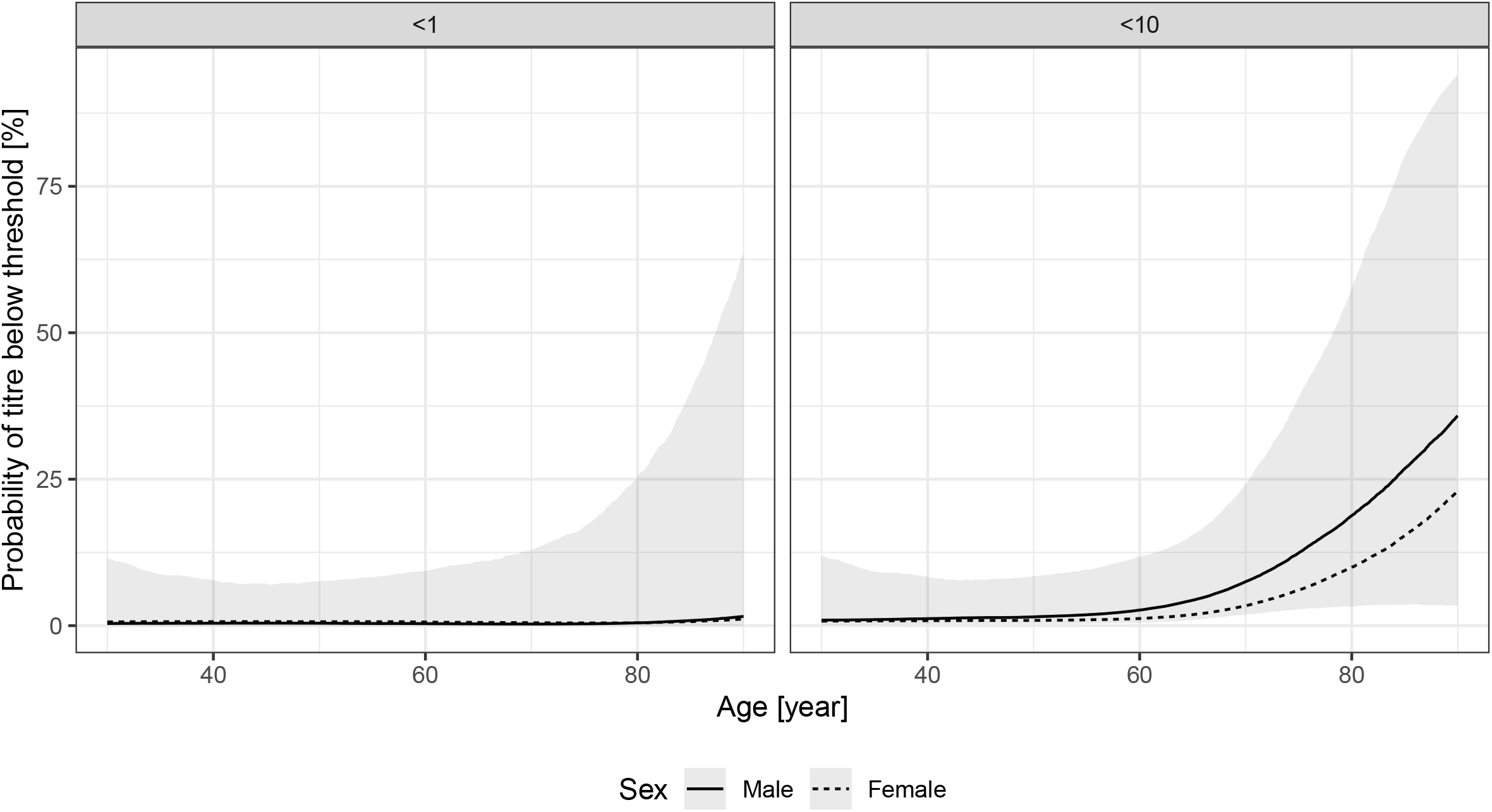
Effects of the age and sex of the subject on the probability of the lack of antibody production (titre below 1, left panel) and of low antibody titre (titre below 10, right panel) after two doses of the Pfizer/BioNTech vaccine. 90% credible interval is shown for males, 28 days post second dose.

## Discussion

In this study we have analyzed the SARS-CoV-2 virus neutralizing antibody titre, as a potential correlate for disease protection, in subjects vaccinated with the Sinopharm inactivated virus vaccine. As to our knowledge, this is the first systematic evaluation of well-defined laboratory measurements obtained from a relatively large number of subjects for the Sinopharm vaccine.

Under the current pandemic circumstances, the reliable estimation of a direct relationship between the risk of the COVID-19 disease, hospitalization or death and the type of vaccine administered, while being the most relevant evidence, may be difficult, especially in low-income countries, and does not deliver individual-level information on susceptibility. However, it is relatively straightforward to analyze already available data on the virus neutralizing antibody titres in the sera of subjects, which, when done with appropriate statistical tools and uniform laboratory methodology, can rapidly generate much-needed evidence assuming its good surrogacy.

The applicability of the virus neutralizing antibody titres, as correlating measures with the COVID-19 disease, has not been fully clarified in the relevant literature^28–32^. While rapid, lateral flow or other IgG-IgM test procedures may yield questionable results, the properly assayed antibody titres have been documented to correlate with disease susceptibility. This is especially true for accredited laboratory assays, as applied here, measuring antibody binding to the receptor binding domain (RBD) of the Spike virus protein, having a key role in the binding and cellular entry of the SARS-CoV-2 virus. This type of assay, that is RBD-binding antibody measurements have been used in the Phase 1-2 studies of the RNA-based vaccines^33^, and has been shown to correlate with direct virus neutralizing assays^24,34^, not available for larger scale studies. Quantification of neutralizing antibody levels is clearly essential for vaccine development and for assessing protective immunity both at the individual and community levels. In addition, RBD binding virus neutralizing antibodies are most promising therapeutic tools for preventing or curing the COVID-19 disease^35,36^.

The major strength of the present study is that it uses a relatively large sample (almost 450 subjects) for the Sinopharm vaccine with a uniform laboratory method and information on age, sex and time since vaccination, entered into a sophisticated statistical model. The sample size allowed the fine exploration of the effect of age (i.e., no categorization was used). To our best knowledge no such study has been published in the scientific literature, despite the fact that data on the protection conferred by the Sinopharm vaccine, and its age-dependence would be crucial for the more than 50 countries where this vaccine is authorized^22^.

The study also has several limitations. First, the sample is not randomly taken from the population (to the contrary, it is likely that participants had a socioeconomic state above average for instance). This, however, is unlikely to have a major impact on antibody response, and especially on its age dependence. Also, while we excluded subjects with known previous SARS-CoV-2 virus infection, asymptomatic infection before vaccination cannot be ruled out. Note that this biases the probability estimate of having low titre after vaccination downwards, so the actual results can be even worse. Third, we had no information on the comorbidities of the subjects, as the study was primarily aimed at assessing the impact of age. Age is correlated with the presence of many chronic diseases, but the effect of these could not be separated in this study. The presented methodology however can be used to rapidly generate evidence in such groups, i.e., subjects with certain comorbidities too. Finally, while accumulating evidence points to the good surrogacy of the antibody titre used in the study, it is nevertheless not a direct measurement of disease susceptibility.

Based on the results presented here, we found that virus neutralizing antibodies were present in a large percentage (about 90%) of individuals below the age of 50 years, while this antibody production after Sinopharm vaccination was much less pronounced with increasing age. In fact, above the age of 60, numerous subjects were found not to produce any protective antibody, reaching a percentage of up to 50% at older ages. When compared to our limited data set of the Pfizer/BioNTech vaccines (the results of which matched those found in major clinical studies^20^), the antibody titres after Sinopharm vaccination were almost an order of magnitude lower than after the Pfizer/BioNTech vaccine; in this latter case we did not find any individuals with no protective antibody.

These findings call for the further evaluation of the protective antibody titres in the large population of elderly people vaccinated with the BBIBP-CorV (Sinopharm, Beijing CNBG) vaccine, and/or to provide additional vaccination of the potentially unprotected individuals. A significant danger is that if the lack of antibody production indeed translates to a weaker protection against the disease, while the Sinopharm vaccinated elderly feel and are declared to be protected against COVID-19, then this false promise may contribute to an outbreak of the disease in this highly vulnerable population.

## Data Availability

De-identified individual patient level data is openly available to the public. Together with the full analysis script it can be found at https://github.com/tamas-ferenci/SinopharmAntibodyResponse.

## Acknowledgment

The authors would like to thank Júlia Singer for her instructive comments that greatly improved the manuscript.

## Statements

### Author contributions

TF carried out the statistical analysis, verified the underlying data, participated in the interpretation of the results and revised the manuscript. BS generated the research question, conceived the study, did the literature review, curated and verified the data, and wrote the manuscript.

### Declaration of interests

TF has no relevant conflict of interest to declare. BS has no relevant conflict of interest to declare.

### Role of the funding source

No specific funding was used to carry out the study.

### Role of medical writer or editor

No medical writer or editor was involved in the creation of the manuscript.

## Notes

### Competing Interest Statement

The authors have declared no competing interest.

### Author Declarations

Used data were openly available to the public before the initiation of the study, as documented in the Data Sharing section.

